# A thematic analysis of Prevention of Future Death Reports for Children who died by suicide in England and Wales: January 2015 to November 2023

**DOI:** 10.1101/2025.05.15.25327671

**Authors:** Emma Sharland, Emma Wallace, Lauren Revie, Isobel Ward, Cathryn Rodway, Daniel Ayoubkhani, Vahé Nafilyan

## Abstract

**Background:** Suicides in children and young people are a major public health concern. Prevention of Future Death (PFD) reports are an underutilised resource detailing coroners’ concerns which if actioned are believed to be able to prevent future deaths. Research has investigated common themes for suicide during 2021 and 2022 but there are no published studies that thematically analyse these reports for children alone.

**Aims:** To identify key themes raised by coroners from PFD reports published between 2015 and 2023 for children who have died by suicide.

**Method:** PFD reports for suicides in children were downloaded from the Courts and Tribunals Judiciary website. Descriptive statistics were collated from reports. Reports (n=37) were analysed using inductive content analysis to determine primary and sub-themes using QSR NVIVO 14 Qualitative Analysis Software.

**Results:** Reports came from 30 coroners’ areas, with most reports being sent to Government Departments and NHS Trusts/Clinical Commissioning Groups. The qualitative analysis resulted in six primary themes being identified: service provision, staffing and resourcing, communication, multiple services involved in care, accessing services and access to harmful content and environment. Furthermore, 23 subthemes were identified such as standard operating procedures/ processes not being followed or inadequate, a lack of specialist services and a disconnect between integrated services. A quarter of reports were in children diagnosed with autism, and there were specific issues highlighted in concerns relating to services and staffing for children with neurodiverse conditions.

**Conclusions:** The key findings from this report highlight themes raised by coroners relating to deaths of children by suicide. This included themes around service provision, staffing and resourcing of mental health services and communication between services and families. There was also a common thread highlighting many concerns relating to children with neurodiversity such as Autism.

## Introduction

The National Suicide Prevention Strategy published in 2023 identified children and young people as a key priority group for targeted and tailored support for suicide prevention^1^. There has been a steady increase year-on-year of suicide in children and young people making this a major public health concern^2,3^. Suicide prevention strategies and policies can only be fully developed when the complex context surrounding suicide in this age-group is understood.

Prevention of Future Death (PFD) reports are written by coroners to raise concerns surrounding a death of a person, where the death was deemed to be preventable^4^. Coroners have a duty under paragraph 7(1) of Schedule 5 of the Coroners and Justice Act 2009 to make PFD reports in order to prevent other deaths from occurring^5^. Research has identified potential benefits for policy makers, health and social care providers, employers and professional bodies in using these reports in research to increase learning and utilisation of them to address repeated issues^6,7^. Inadequate processes and documentation, and communication concerns, were some themes identified from PFD reports for suicide (all ages) published between January 2021 and October 2022^8^. However, PFD reports have not previously been used to identify concerns and areas for prevention specifically for suicides in children and young people. Previous research using other data sources has identified common themes relating to suicide in children and young people, including academic pressures, social isolation, and mental ill health and self-harm^2,9^.

The aim of this study was to build on previous research by analysing PFD for suicides in children. The analysis of these reports will allow for themes and patterns of failures to be identified and will provide clear evidence for policy makers when developing future interventions and support for children and young people who are at risk of suicide.

## Method

### Study Design and Data

We used a retrospective case-series design. PFD reports, which are publicly available on the Courts and Tribunals Judiciary website, were used in the analysis^4^. These reports are completed for deaths where a coroner felt the death could have been prevented and action could be taken to prevent future deaths. Reports are available from 2013 in PDF format and cover both England and Wales. Reports are generally categorised by type of death, for example, “Suicides (from 2015)”, “Child death (from 2015)” and “Care Home related death”. The categories for suicides and child deaths were introduced in 2015. All reports typically have a standard structure including a section detailing the circumstances of the death and coroner’s concerns. There are occasions where parts of the reports are redacted, and they therefore could not be included in our analysis. Reports are often addressed to one or more people or organisations, who the coroner believes should act upon the concerns raised in the report.

The authors assert that all procedures contributing to this work comply with the ethical standards of the relevant national and institutional committees on human experimentation and with the <u>Helsinki Declaration</u> of 1975, as revised in 2013. All procedures involving human subjects/patients were approved by the National Statistician’s Data Ethics Advisory Committee (*NSDEC(22(17))*. Consent was not obtained from participants in this study due to PFD reports being freely available on the Courts and Tribunals Judiciary website; this approach was approved by the NSDEC ethics committee.

### Patient and Public Involvement

We engaged with adults who had lost a child or family member to suicide, or who were supporting children or family members with suicidality either in a personal or professional capacity. This engagement was to discuss the themes emerging from the analysis, to understand how they related to lived experiences. We also had a paper review group who came from the University of Manchester’s Centre for Mental Health and Safety central PPIE group. Members of PPIE groups were involved in reviewing the results of the study and the reporting of it. All PPIE reviewers also contributed to the dissemination plans for this research.

### Procedure

PFD reports categorised as “Child Death (from 2015)” with an additional category of “Suicide (from 2015)” and/or “Mental Health Related Death” were reviewed for analysis in January 2024. Reports categorised as “Child Death (from 2015)” and “Mental Health Related Death” were only included in analysis if the death was from suicide, which was identified from the content of the report. As reports can have multiple categories applied, the reports analysed in this study also included other categorisations such as “Railway related death”. Child deaths are defined on the PFD reports as a person aged 18 years and under at the time of death. Reports included in the analysis were published on the Courts and Tribunals Judiciary Service website between 1^st^ January 2015 and 30^th^ November 2023. Reports (n=37) were manually downloaded for analysis. Three reports published on the website related to two individuals: one individual report each and a combined report. All three reports were included in the analysis as the information and concerns raised were distinct in each of the reports and there was no concern of duplication. As of March 2024, 319 reports were classified as a child death out of a total of 4204 PFD reports (from 1^st^ January 2015). Therefore, the analysis covered 12% of child death reports and 0.9% of PFD reports overall.

We conducted descriptive analysis on the number of concerns per report (total, mean and range)^6,8^. Additional counts and percentages for sex, age and number of recipients of the report were recorded. Concerns were coded into primary and sub-themes (to provide additional context), and concerns raised could cover multiple primary and sub-themes if relevant. Concerns were identified from the “Coroner’s Concerns” section of the PFD report. Additionally, concerns were analysed if found within the “Circumstances of the death” section of the reports, where a researcher felt the issue raised related to a concern about the circumstances of the death. All suicides are subject to a coroner’s inquest, and in 2023 the median registration delay was 199 days in England and 293 days in Wales^3^; therefore, the publication of a PFD report can be significantly later than the date that the death occurred.

All PFD reports (n=37) were analysed in QSR NVivo 14 Qualitative Analysis software by one experienced researcher (EW) initially. They used an inductive content analysis approach to identify common themes within the “Coroner’s Concerns” and “Circumstances of the death” sections of the reports. This same researcher then reviewed the coding structure to refine the codes further and produce the primary and sub-themes. A secondary researcher (LR) also analysed all PFD reports. The initial agreement between the two researchers was 66% ((Agreed codes ÷ Total number of codes) × 100 = Agreement (%)). Following a review of the codes within the primary and secondary themes, agreement between the two researchers was reached for 99% of codes. For the remaining 1% of codes, the lead researcher’s decisions were used.

## Results

### Summary of the Study Data

A total of 37 PFD reports were descriptively and thematically analysed. A total of 145 concerns were raised across these PFD reports with an average of 4 concerns per report (range 1-12). The average number of days between the date of death and the publication of the PFD report was 626 days (range 152-1,788 days). Reports covered deaths occurring between 2012 and 2022.

All PFD reports in this analysis were categorised as a “Child death (from 2015)”, and all reports also had either “Suicide (from 2015)” (76%) and/or “Mental health related deaths” (62%) categorisations (Table 1). Other categorisations included in the analysed reports are shown in Table 1.

**Table 1:** Categorisation of PFD reports used in this analysis. “Child death (from 2015)” is not included as all reports included in the study (n=37) had this category.

| Categorisation | Count |
| --- | --- |
| Suicide (from 2015) | 28 |
| Mental Health related deaths | 23 |
| Hospital death | 11 |
| Other related deaths | 8 |
| Railway related deaths | 6 |
| Alcohol, drug and medication related deaths | 5 |
| Other<sup>1</sup> | 7 |

<sup>1</sup>Other includes the categories: “Police related deaths”, “Community health care and emergency services related deaths” and “Care Home related deaths”.

Reports came from 30 of the 83 coroners’ areas in England and Wales (see limitations section). PFD reports can be addressed to multiple people, organisations, and agencies. One or more government departments or ministers were addressees in almost half of the PFD reports (n =15, 41%), as were NHS Trusts and Clinical Commissioning Groups (CCGs) (n=15, 41%) (Table 2).

**Table 2:** Addressees of PFD reports (n=37).

| Addressee | No. of reports | % |
| --- | --- | --- |
| Government department or minister | 15 | 41 |
| NHS Trust or CCG | 15 | 41 |
| Professional body | 12 | 32 |
| Local council | 8 | 22 |
| Other<sup>2</sup> | 10 | 27 |

<sup>1</sup>PFD reports can be addressed to multiple organisations so total will not equal 37. <sup>2</sup> “Other” is used where numbers are less than 3 and therefore disclosive. “Other” includes, rail or road company, police (including British Transport Police), private companies and hospital or Child and Adolescent Mental Health Services (CAMHS).

### Descriptive Statistics

Of the 36 reports^1^, 10 (27%) did not specify the age of the deceased. Where age was provided for the deceased, the average age was 16 years (range 12-18 years). Most reports related to females (n = 19, 53%) and 44% (n = 16) of reports related to males. Sex was only reported in 97% of the reports.

Of the 36 PFD reports, 32 (88%) detailed diagnoses of the deceased where it was relevant to the report. Aspergers syndrome or autism spectrum disorder was reported in a quarter of reports (25%), followed by mood disorders (19%) and anxiety disorders (14%) (Table 3).

**Table 3:**
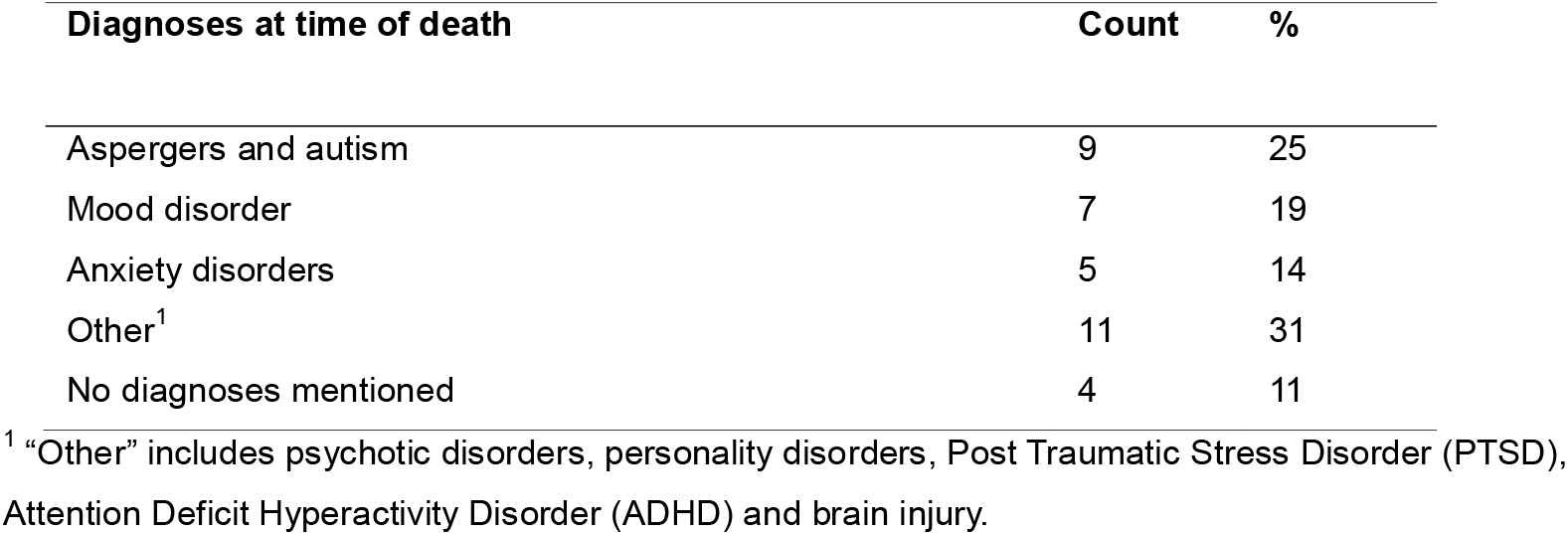
Diagnoses mentioned on PFD reports (n=36).

| Diagnoses at time of death | Count | % |
| --- | --- | --- |
| Aspergers and autism | 9 | 25 |
| Mood disorder | 7 | 19 |
| Anxiety disorders | 5 | 14 |
| Other<sup>1</sup> | 11 | 31 |
| No diagnoses mentioned | 4 | 11 |

<sup>1</sup> "Other" includes psychotic disorders, personality disorders, Post Traumatic Stress Disorder (PTSD), Attention Deficit Hyperactivity Disorder (ADHD) and brain injury.

11% (n = 4) of reports indicated the deceased had a history of detention under the mental health act, 64% (n = 23) were known to CAMHS, 39% (n = 14) had a history of self-harm and 42% (n = 15) indicated a history of previous suicide attempts or suicidal thoughts.

### Thematic Analysis

The thematic analysis identified six primary themes and 23 sub-themes (Table 4). For details on sub-themes, see supplementary taable 1.

**Table 4:** Primary and sub-themes identified from PFD reports (n=37) and number of mentions of each sub-theme.

| Primary theme | Sub-theme | Count |
| --- | --- | --- |
| Service provision | Standard operating procedures/ processes not followed or inadequate | 40 |
|  | Lack of specialist services or support (crisis, autism, beds) | 22 |
|  | Risk assessment not completed or reviewed<sup>1</sup> | 20 |
|  | Discharges from services were ineffective | 8 |
|  | Lack of support for children with specific diagnoses | 4 |
| Staffing and resourcing | Training missing, inadequate or not mandatory | 17 |

<sup>1</sup> One report was removed from descriptives due to it being a joint report between two deceased people who also have individual reports.

|  |  |  |
| --- | --- | --- |
|  | Inadequate staffing | 10 |
|  | Lack of funding | 10 |
|  | Recruitment and retention problems | [c] |
| Communication | Lack of communication between services | 19 |
|  | Lack of communication with patient and family | 9 |
|  | Confidentiality risk not communicated | [c] |
|  | Within services communication is poor | [c] |
| Multiple services involved in care | Integration of care was disconnected | 16 |
|  | Issues with Local Authority (including child services, schools) | 11 |
|  | Transition from CAMHS to adult services ineffective | 4 |
| Accessing services | Delays in referrals and waiting times | 13 |
|  | Referrals rejected | 5 |
|  | Patient engagement lacking | 5 |
| Access to harmful content and environment | Internet content and controls | 9 |
|  | Safeguarding from sensitive material | 5 |
|  | Access to harmful items/ substances | 4 |
|  | Access to Trainlines | [c]<sup>2</sup> |

<sup>1</sup>Since September 2022, National Institute for Health and Care Excellence (NICE) guidelines state that risk assessment tools should not be used in the prediction of future suicide or repetition of self-harm. Themes relating to risk assessment are included in this analysis because of the clinical relevance of risk assessments at the time these reports were written. <sup>2</sup>Where number of mentions shows confidential [c], this result is below 3 and therefore potentially disclosive.

#### Service provision

Service provision was the most common primary theme and had five sub-themes. The main sub-theme related to standard operating procedures or processes not being followed or were inadequate (n=40), including missing processes/protocols, lack of escalation, and ineffective monitoring and reporting. The second most frequent sub-theme related to a lack of specialist services (n=22). The details of this theme included lack of available services, inappropriate support with children being placed in adult care services, and no case workers or key workers assigned for children with specialist needs such as autism. For specialist services, the reports detailed unavailable beds in urgent inpatient care, crisis teams out of hours and specialist services for children with autism.

#### Staffing and resourcing

The second most common primary theme related to staffing and resourcing. The main sub-theme related to training of staff (n=17). Reports detailed that either staff were not mandated to complete certain training, training did not exist, or training was ineffective. Specific examples related to children with additional needs such as autism, where there was a lack of mandatory training for those working specifically with children with these needs, despite a working knowledge of links to mental ill-health or suicidal thoughts. Also, inadequate staff knowledge of suicide prevention processes and gaps in knowledge of standard operating procedures. Inadequate staffing (n=10) included issues where there were no staff available and staff not having the appropriate qualifications to meet the needs of the child involved.

There were also mentions of staff managing high caseloads or children experiencing long delays due to lack of qualified staffing. Lack of funding was highlighted as a sub-theme where reports raised concerns about under resourcing of services and funding for schemes being the same year on year, despite referrals increasing substantially. Reports suggested that ultimately, these issues led to resources not being able to meet demand and consequently putting lives at risk.

#### Communication

Within the primary theme of communication, communication between services was reported to be lacking (n=19). This captured communication between different health teams or between other organisations such as transport and emergency services. The communication issues between services led to both information not being passed on to support children and families, and a lack of risk being highlighted to other services where mitigating factors such as additional support or monitoring could have been implemented. A lack of communication with the family of the individuals prior to their death was also raised as a concern (n=9).

Reports identified additional information and context that may have been given by the family which could have helped support an individual and reduce the risk of suicide.

#### Multiple services involved in care

A disconnect between integrated services (n=16) involved in the care of an individual was identified within reports raising issues within organisations (multiple departments) and between organisations. The reports suggested that care became ineffective, and individuals’ needs and support (including when there was history of mental ill-health and suicide risk or additional needs due to disabilities) were not adequately met. Issues with local authority care (n=11) were also reported, with areas not effectively reviewing the circumstances of a death leading to limited learning following a suicide event and no mitigation of future risk by organisational changes.

#### Accessing services

The main sub-theme that emerged under the primary theme of accessing services was delays in referrals being made and long waiting times (n=13). The concerns raised in the reports suggested that waiting times were too long and some waits were due to bed shortages in services such as CAMHS and specialist mental health services. Additionally, delays in receiving diagnoses for conditions such as autism as well as access to specialist services for these conditions reportedly resulted in individuals not receiving support or receiving inappropriate support for their needs.

#### Access to harmful content and environment

The internet was the main sub-theme identified for the access to harmful content and environment (n=9) theme. Within this subtheme there were issues raised around the lack of parental controls available (including control the over the content, for example, age verifications and links between parent and child accounts), social media use and the type of content being viewed being inappropriate and individuals coming across content encouraging suicide and potential methods.

## Discussion

This study is the first to explore themes from qualitative analysis of concerns raised by coroners in PFD reports in England and Wales for children and young people who died by suicide. The main theme emerging from the research was around lack of service provision, followed by inadequate staffing and breakdown in channels of communication.

The primary theme of service provision focussed on processes not being followed or being inadequate, and the lack of provision of services including specialist services. Similar to Wallace et al. (2024)^8^ study of PFD reports for suicide across the population processes like standard operating procedures not being followed were a prominent concern for suicides across the population, indicating a wider issue with mental health and suicide prevention services. The National Suicide Prevention Strategy, published in the 2023, aims to improve information sharing and access to timely and effective crisis support in England^1^. The issues of staffing raised in the present work are corroborated across other research into PFDs^8,10^. Several studies suggest that a lack of funding and an increase in demand for children and young peoples mental health services is resulting in problems in accessing appropriate support, including staffing shortages (particularly for psychiatrists and mental health nurses) and training existing staff and this is resulting in delays to referrals and admissions, long waiting times, and receiving care in inappropriate settings^11–13^. Following an expansion of Alder Hey’s children’s crisis care line a 50% reduction in the number of hospital admissions was observed^14^. Additional evaluation would be required to assess if this is an effective intervention in other regions.

Previous research has highlighted a potential increased risk of suicide amongst people with autism or autistic traits. A study investigating the association of autism diagnoses and autistics traits and suicidal ideations and self-harm in adolescents found an association between impaired social communication and increased risk of suicidal thoughts, plans and self-harm with suicidal intent^15^. These children have a higher risk of depressive symptoms in early adolescence and the authors emphasised the need to address mental health needs in children with autism^[14]^. Recent research in adults has also found around 10.7% of suicides in England were among people with autism or those who had autistic traits (11 times higher than the prevalence of autism in the UK), and this was a likely underestimate^16^. Research also reports that there has been an increase from 17 per year (2011-2014) to 49 per year (2018-2021) of autistic people dying by suicide who were patients of mental health services^17^. An autism diagnosis was reported in 25% of the reports in the present study and many of the primary and sub-themes involved concerns relating to the support given to children and young people with autism. Additionally research conducted on PFD reports for people with Autism found 67% had died by suicide^18^. Cassidy et al. (2022)^16^ recommended screening for autism more widely in people who have suicidal thoughts or mental-ill health to ensure the correct support is in place. Concerns around the delays in diagnoses of autism were also raised in this study. There is a growing backlog of adults and children in England who are waiting for an autism diagnosis, with some 172,022 patients on the waiting list in December 2023 and 85.5% waiting for over 13 weeks, longer than the wait time recommended by the National Institute for Health and Care Excellence (NICE)^19,20^. Delayed diagnoses or undiagnosed autism are reported to potentially be preventable suicide risk factors^1^. Autistic people often report feeling excluded from services and often receive inappropriate support including a lack of specialised provision and treatment^1,16,18^. The findings from our study support this research, with concerns raised around a lack of specialised support for autistic children and a lack of suitably qualified or trained staff who know how to appropriately support these children. It is recommended in the Governments Suicide Prevention Strategy and by Cassidy et al (2022) that reducing barriers to accessing appropriate support, including reducing waiting times and giving increased consideration for the needs of autistic people, should be routinely included in suicide prevention activities^1,16^.

Communication between services and with the families of the individuals was a key theme raised within this study, as per previous research using concerns from PFDs^6,8,10^. A previous systematic review identified the need for improved communication between primary care and mental health services as essential throughout the care process for people who self-harm^21^. Research has identified that a person’s next of kin or family members have useful information to support the care of an individual with previous suicide attempts, suicidal ideations or in mental health crises care^22,23^. In a study in Ireland looking at parental experiences with CAMHS services, some parents felt their involvement was suboptimal, with parents explaining they felt they had to fight for their voice to be heard, or to get information on the progress of their child in the service^24^. Parents also felt uncomfortable speaking with health care providers about personal issues without judgement, and that their children did not feel they could talk openly^24^. Supporting the primary theme of staffing established in this study, research has also identified that staff turnover in CAMHS negatively impacts communication between the families and the service, with parents feeling like this resulted in stories having to be re-told^24^. The literature highlights that communication with families plays an important role in caring for people who are undergoing a mental health crisis^21,23^, and it should be part of standard processes for care providers to gather as much information as possible, and keep communication non-judgemental, to understand the individual they are caring for as per NICE guidelines^25^.

Despite previous research indicating that a quarter of young people have used the internet to research suicide methods, communicate suicidal thoughts and seek encouragement^26^, concerns relating to internet controls and content were not prominent within coroners’ reports when compared to processes, staffing and communication issues. The Online Safety Act looks to promote online safety and responsible content, recommends consistent and responsible portrayal of self-harm and suicide content and safer platforms for use^1^.

### Limitations

The number of reports used in this analysis was small (n=37) compared to the total number of suicides among children registered annually^3^. In 2023, there were 219 suicides registered in 10 to 19 year olds in England and Wales and, across the period of 2012 to 2023 there were 2,197 suicides registered in the same group^3^. This is due to PFD reports only being written for a select few deaths where the coroner had concerns to raise to prevent a death in the future. Therefore, the themes identified from this analysis may not be generalisable to all deaths in children from suicide. Importantly, the key themes identified in our analysis are restricted to the content published in the PFD reports and therefore are not exhaustive of all potential issues related to suicide prevention. For instance, online safety was not identified as a prominent factor in our analysis, however previous research^27^ and our patient and public engagement suggest this as a key area for future suicide prevention. The lack of evidence emerging from this study for other factors does not mean these factors are not important.

In this study we have conducted analysis on reports categorised on the Courts and Tribunals Judiciary website as a “Child death (from 2015)” and “Suicide (from 2015)” and/or “Mental health related death”. The categorisations for child death and suicide were only introduced in 2015, therefore there may be reports published prior to 2015 that have not been included in the study. Additionally, there may be relevant reports where only one or no categorisations have been applied and were therefore not included. Research analysing the PFDs uploaded to the Courts and Tribunals Judiciary website found around 33% of reports in 2021 had not been categorised^27^. Some studies have indicated limitated application of categorisation of reports for suicide with around 19% of reports categorised correctly, so some relevant reports may have been excluded from the analysis^28^. Additionally, age is not always noted on reports, therefore there may have been reports relating to a child death which have not be categorised as such and therefore excluded.

The COVID-19 pandemic is likely to have had an impact on the publication of PFD reports, due to the long delays between a death occurring and the publication of the report. It is possible that in the coming years more PFD reports will be published covering the pandemic period, which may raise different concerns and themes not included in this study.

Research has found that the top 20 coroners who complete PFD reports account for 30% of reports on the website^27^. This means the reports analysed in this study may be biased towards those coroners who are more likely to complete a PFD report and subsequently the areas that they work within.

### Conclusions

This study has identified key themes from PFD reports for children and young people who have died by suicide where coroners have raised concerns and where a death could have been prevented. This included themes around service provision, staffing and resourcing of mental health services and communication between services and families. There was also a common thread throughout highlighting many concerns relating to children who died by suicide with neurodiversity such as autism which may be an important group for further research into suicide prevention.

## Data Availability

Data Availability: Prevention of Future Death reports are made publicly available at https://www.judiciary.uk/courts-and-tribunals/coroners-courts/reports-to-prevent-future-deaths/. Accompanying datasets for this publication can also be found at the ONS website.

## Ethics Statement

This project was approved by the National Statistician’s Data Ethics Advisory Committee (*NSDEC(22(17))*

## Acknowledgements

The authors would like to thank David Mais (Office for National Statistics) for his contributions to reviewing the analysis and in the Patient and Public Involvement and Engagement.

We would like to thank the participants of our two public and patient advisory groups. Our groups comprised of young adults with experiences of suicidality or supporting others with suicidality, and secondly adults who had lost a child or family member to suicide, or who were supporting children or family members with suicidality either in a personal or professional capacity. Members of our public and patient advisory groups reviewed the results of the analysis and provided feedback. We would like to thank Papyrus UK and The Mix for facilitating these workshops.

We would also like to thank members of the Mutual Support for Mental Health Research (MS4MH-R) Patient and Public Involvement and Engagement group affiliated with the NIHR Greater Manchester Patient Safety Research Collaboration and Centre for Mental Health and Safety at The University of Manchester for their contribution to the interpretation of results and reviewing of publications.

## Author Contribution

ES and IW conceived and designed the study. EW and LR analysed, and quality assured the data. ES and IW wrote the manuscript, with contributions from CR, DA and VN. The corresponding author attests that all listed authors meet authorship criteria and no others meeting the criteria have been omitted.

## Transparency Declaration

The first author (ES) is the guarantor and affirms that this manuscript is an honest, accurate and transparent account of the study being reported; no aspects of this study have been omitted; and that any discrepancies from the study as planned have been explained

## Data Availability

Prevention of Future Death reports are made publicly available at https://www.judiciary.uk/courts-and-tribunals/coroners-courts/reports-to-prevent-future-deaths/. Accompanying datasets for this publication can also be found at the ONS website.

## Analytical code availability

Not applicable

## Research material availability

Materials supporting the findings will not be publicly available to other researchers.

