## Supplementary table 1 for "A thematic analysis of Prevention of Future Death Reports for Children who died by suicide in England and Wales: January 2015 to November 2023"

**Supplementary table 1: Definitions of primary and sub-themes**

| **Main theme** | **Subtheme** | **Description of subtheme contents from concerns arising in the reports** |
| --- | --- | --- |
| **Service provision** | Specialist services (crisis, autism, beds) | Specialist urgent inpatient beds not available (i.e. on a child psychiatric ward) for children during time of crisis |
|  |  | Specialist urgent crisis team (out of hours support) not available for children being looked after in the community |
|  |  | Special Educational Needs / Autism support services don't exist or unavailable in catchment areas |
|  |  | Due to the care requirements of longer-term conditions (for example, Autism), some evidence suggested that the structure of Clinical Commissioning Groups/ Trusts biases against these conditions and prioritisation is not given to related specialist services |
|  | Discharge from services | Discharge failed to involve a review or liaison officer/ individual |
|  |  | Individual self-discharged where Mental Health Act / detainment may have been required for safety |
|  |  | Care requirements or risk of the individual not communicated to community teams upon discharge |
|  |  | Care not coordinated when discharged from crisis care |
|  |  | Care package did not meet individuals needs or failed to cover all instances (e.g. items or environments which could trigger an event) |
|  | Standard operating procedures/ processes | Standard operating procedures (e.g. note taking, monitoring or observations) not followed correctly by service staff |
|  |  | Standard operating procedures don't exist or are unclear |
|  | Diagnostics | Delay in correct diagnosis |
|  |  | Misdiagnosis |
|  |  | Lack of support related to the care of individuals with a specific diagnosis (e.g. autism) for caregivers |
|  |  | Specialist diagnosis training for staff not available |
|  | Risk assessment | No risk assessment documents completed |
|  |  | Level of risk assessed was inadequate |
|  |  | Risk assessment wasn't updated |
|  |  | Risk wasn't fully communicated to staff |
| **Staffing and resourcing** | Inadequate staffing | Staff not appropriately qualified (e.g. experienced psychiatrists) |
|  |  | Inexperienced case worker assigned to the case, resulting in gaps in service provision |
|  | Training | Inadequate staff knowledge of suicide prevention processes |
|  |  | Inadequate provision / preparation for staff (e.g. no presence of grab bags or anti-ligature tools where required) |
|  |  | Staff not following Standard Operating Procedures due to gaps in training knowledge |
|  | Funding | Lack of funding to Child and Adolescent Mental Health Services (CAMHS) services meaning staff cannot be recruited |
|  |  | Lack of funding meaning specialist services cannot be provided |
|  | Recruitment and retention | Unable to recruit specialist staff |
|  |  | Service unable to retain adequate amount of staff |
| **Communication** | Between Services | Lack of communication between Child and Adolescent Mental Health Services (CAMHS) and foster/ care services |
|  |  | Lack of communication between Child and Adolescent Mental Health Services (CAMHS) and schools |
|  |  | Information sharing between services (e.g. medical history / care) not possible or not conducted |
|  | With patient and family | Lack of communication from Child and Adolescent Mental Health Services (CAMHS) with child and/or parent |
|  |  | Lack of family involvement in care / lack of support or signposting from services for the family |
|  | Within services | Inadequate communication of policies to staff |
|  |  | Inadequate note keeping and/or record sharing |
|  |  | Lack of communication within services regarding who is the care coordinator / responsible party for joined up care provision |
|  | Confidentiality risk not communicated | Instances where professional could have, but did not communicate with parents or caregiver due to patient confidentiality, resulting in missed opportunity to engage parents or communicate risk |
| **Multiple services involved in care** | Integration of care | Care coordinator not assigned or unclear who was coordinating individuals care needs |
|  | Transition from CAMHS | Lack of support to the individual when transitioning from Child and Adolescent Mental Health Services (CAMHS) to adult services |
|  |  | Guidance on patient qualification for Adult vs Child and Adolescent Mental Health Services (CAMHS) service (e.g. where the individual is 16 to 18 yrs) unclear |
|  | Local Authority (including child services, schools) | Lack of social services involvement |
|  |  | No social worker assigned |
|  |  | Inadequate safeguarding checks |
|  |  | Lack of specialist support staff in schools (e.g. Special Educational Needs / counsellor) |
|  |  | Safety plan not present in schools |
| **Accessing services** | Delays in referrals and waiting times | Delay in referral from General Practitioner and Child and Adolescent Mental Health Services (CAMHS) |
|  |  | Delay in Child and Adolescent Mental Health Services (CAMHS) team picking up referral |
|  |  | Delays in Child and Adolescent Mental Health Services (CAMHS) / specialist mental health service offering appointment |
|  |  | Excessive waiting times leading to General Practitioner / medical professional making inappropriate referral |
|  |  | Covid-19 related delays in referral and treatment |
|  | Referral rejected | Referral to Child and Adolescent Mental Health Services (CAMHS) rejected due to waiting times / lack of staff / risk not adequately assessed for the individual |
|  |  | Complex needs of the patient can't be met by Child and Adolescent Mental Health Services (CAMHS) |
|  | Patient engagement lacking | Inadequate contact with child or adult regarding referral (e.g. no follow up call to offer alternative appointment) |
|  |  | Patient refusal to engage (Child and Adolescent Mental Health Services (CAMHS) subsequently not following up with parent or caregiver in proactive manner) |
| **Access to harmful content and environments** | Internet | Lack of internet safeguarding either in school or failure of websites / social media to block harmful content |
|  | Trainline | Ability to access the trainline where access should not be permitted / available (e.g. inadequate fencing) |
|  | Harmful items/ substances | Access to items that can be used to harm or ligature, where it was known these are a trigger or there is a safety concern |
|  |  | Access to alcohol / drugs / substances where a safety concern is known surrounding these substances |
|  | Safeguarding from sensitive material | Sensitive questions asked of the child or presented to the child without adequate follow up or adult support |
|  |  | Sensitive material presented to the child in school (as part of the curriculum) without precautions / warnings / consideration of child safety |
| **Place of death** | Hospital (inpatient psychiatric) | Place of death was at an inpatient psychiatric ward (adult or child) where the individual had been admitted and was receiving treatment |
|  | Hospital (general) | Place of death was at hospital following an emergency admission / was not related to a psychiatric inpatient stay |
|  | Railways and road network | Place of death was a trainline, underground station, highway or motorway |
|  | Outdoor spaces | Place of death was a park, quarry, field, cliff, or other outdoor space |
|  | Home address (private) | Place of death was the deceased's known family home address (parent, foster carer, family member) |
|  | Home address (communal establishment) | Place of death was a communal establishment (supported living, children’s home, shelter, crisis centre) where the child was residing |
| **Diagnosis at time of death** | Post Traumatic Stress Disorder (PTSD) | Including post-traumatic stress disorder |
|  | Psychotic disorder | Including schizophrenia, unknown psychosis, schizoaffective disorder |
|  | Mood disorder | Including depression |
|  | Anxiety disorders | Including generalised anxiety disorder, Obsessive Compulsive Disorder |
|  | Brain injury | Including brain injuries following other neurological disorders |
|  | Attention Deficit Hyperactivity Disorder (ADHD) | Including Attention Deficit syndrome and Attention Deficit Hyperactivity Disorder (ADHD) |
|  | Personality disorders | Including borderline or unstable personality disorder |
|  | Autistic Spectrum Disorders | Including Autism, Aspergers |
| **Addressee** | Child and Adolescent Mental Health Service (CAMHS) | Recipient of the Prevention of Future Death report was the Child and Adolescent Mental Health Service (CAMHS) |
|  | Hospital | Recipient of the Prevention of Future Death report was a hospital |
|  | NHS Trust/Clinical Commissioning Group (CCG) | Recipient of the Prevention of Future Death report was an NHS Trust or Clinical Commissioning Group (CCG) |
|  | Community health service | Recipient of the Prevention of Future Death report was a Community Health Service |
|  | Social services | Recipient of the Prevention of Future Death report was Social Services |
|  | Local council | Recipient of the Prevention of Future Death report was the Local Council |
|  | Police | Recipient of the Prevention of Future Death report was the police, including transport police |
|  | Private company | Recipient of the Prevention of Future Death report was a private company, including internet, health, social care/housing |
